# Beliefs associated with Intentions of Non-Physician Healthcare Workers to Receive the COVID-19 Vaccine in Ontario, Canada

**DOI:** 10.1101/2021.02.19.21251936

**Authors:** Laura Desveaux, Rachel D. Savage, Mina Tadrous, Natasha Kithulegoda, Kelly Thai, Nathan M. Stall, Noah M. Ivers

## Abstract

Achieving herd immunity of SARS-CoV-2 through vaccines will require a concerted effort to understand and address barriers to vaccine uptake. We conducted a web-based survey of non-physician HCWs, informed by the Theoretical Domains Framework, measuring intention to vaccinate, beliefs and sources of influence relating to the COVID-19 vaccines, and sociodemographic characteristics. Vaccination non-intent was associated with beliefs that vaccination was not required because of good health, lower confidence that the COVID-19 vaccine would protect their family and patients, and that getting vaccinated was a professional responsibility. Vaccination non-intent was strongly associated with mistrust about how fast the vaccines were developed and vaccine safety concerns. Communication directed at non-physician HCWs should be tailored by ethnic subgroups and settings to increase salience. Messaging should leverage emotions (e.g., pride, hope, fear) to capture interest, while addressing safety concerns and confirming the low risk of side effects in contrast to the substantial morbidity and mortality of COVID-19. Emergent data about reduced transmission post-vaccination will be helpful.

## Background

Achieving herd immunity of SARS-CoV-2 through vaccines will require a concerted effort to understand and address barriers to vaccine uptake. Initial vaccination of healthcare workers (HCWs) is essential because of their high risk of SARS-CoV-2 exposure, close contact with vulnerable populations, and potential to act as role models and influencers^1^. Unfortunately, 1/6 of HCWs are reluctant to receive the COVID-19 vaccine, citing concerns around vaccine safety and effectiveness, data transparency, and coercion^2^.

## Methods

We conducted a web-based survey of non-physician HCWs, informed by the Theoretical Domains Framework^3^, measuring intention to vaccinate, beliefs and sources of influence relating to the COVID-19 vaccines, and sociodemographic characteristics. Vaccination intention was measured with the question “How likely is it that you would get a COVID-19 vaccine?”. Sociodemographic characteristics collected included: age, gender, ethnicity, education, healthcare worker role and setting. Group comparisons were made using χ^2^ tests and corresponding adjusted associations were made using multivariable logistic regression. The survey was distributed electronically via a large Canadian healthcare union in Ontario, Canada from January 4-12, 2021. The study was approved by the Women’s College Hospital Research Ethics Board and reporting followed the Checklist for Reporting Results of Internet E-Surveys.

## Results

The participation rate was 85% (8,634 completed/10,193 accessed). Participants were mostly female (89.2%), healthcare aid/personal support workers (42.8%), employed full-time (67.2%), and working in the continuing care setting (64.6%), which includes home care, supportive living, and nursing homes. Overall, 80.4% of HCWs reported that they were likely to get a COVID-19 vaccine (Table 1).

**Table 1.** Characteristics of participants, overall and by initial willingness to get a COVID-19 vaccine.

| Characteristic | Overall<br>(N=8634 )<br>No. (%) | Intent to vaccinate (n=8298) |  | P-value |
| --- | --- | --- | --- | --- |
|  |  | Likely<br>No. (%) | Unlikely<br>No. (%) |  |
| Age group, years (n=7262) |  |  |  |  |
| 18-29 | 692 (9.5) | 530 (9.0) | 162 (11.9) | <0.001 |
| 30-39 | 1514 (20.9) | 1136 (19.2) | 378 (27.9) |  |
| 40-49 | 1970 (27.1) | 1589 (26.9) | 381 (28.1) |  |
| 50+ | 3086 (42.5) | 2650 (44.89) | 436 (32.2) |  |
| Gender (n=7226) |  |  |  |  |
| Female | 6446 (89.2) | 5206 (88.4) | 1240 (92.8) | <0.001 |
| Male | 780 (10.8) | 684 (11.6) | 96 (7.2) |  |
| Country of birth (n=7176) |  |  |  |  |
| Outside Canada | 2651 (36.9) | 2287 (39.0) | 364 (27.7) | <0.001 |
| Canada | 4525 (63.1) | 3575 (61.0) | 950 (72.3) |  |
| Ethnicity (n=5619) |  |  |  |  |
| Filipino | 861 (15.3) | 781 (16.7) | 80 (8.5) | <0.001 |
| Caribbean | 507 (9.0) | 364 (7.8) | 143 (15.2) |  |
| Other<sup>a</sup> | 2555 (45.5) | 2105 (45.0) | 450 (47.7) |  |
| European | 1696 (30.2) | 1426 (30.5) | 270 (28.6) |  |
| Highest education level (n=7016) |  |  |  |  |
| No certificate, diploma or degree | 60 (0.9) | 45 (0.8) | 15 (1.2) | <0.001 |
| High school diploma | 858 (12.2) | 727 (12.7) | 131 (10.2) |  |
| College diploma | 4753 (67.8) | 3791 (66.2) | 962 (74.8) |  |
| University degree | 1345 (19.2) | 1167 (20.4) | 178 (13.8) |  |
| Healthcare role (n=7211) |  |  |  |  |
| Support workers | 3085 (42.8) | 2459 (41.9) | 626 (46.8) | 0.003 |
| Staff | 1872 (26.0) | 1547 (26.3) | 325 (24.3) |  |
| Other | 698 (9.7) | 563 (9.6) | 135 (10.1) |  |
| Nurse | 1556 (21.6) | 1305 (22.2) | 251 (18.8) |  |
| Setting of employment (n=7124) |  |  |  |  |
| Continuing care | 4605 (64.6) | 3727 (64.2) | 878 (66.4) | 0.140 |
| Clinic/acute care facility | 2519 (35.4) | 2075 (35.8) | 444 (33.6) |  |
| Position (n=7199) |  |  |  |  |
| Part-time/casual | 2358 (32.8) | 1880 (32.0) | 478 (36.3) | 0.003 |
| Full-time | 4841 (67.2) | 4001 (68.0) | 840 (63.7) |  |

<sup>a</sup> Grouped into the 'Other' category were the following ethnicity categories: First Nations, Indigenous, Inuit, or Metis; Latin, Central and South American (e.g. Mexican, Argentinian, Guatemalan); African (e.g. South African, Ethiopian, Nigerian); Arab/West Asian (e.g. Lebanese, Moroccan, Iranian, Turkish); South Asian (e.g. East Indian, Pakistani, Sri Lankan, Bangladeshi); Other East and South East Asian (e.g. Vietnamese, Korean, Japanese); or Other

After adjustment (Table 2), sociodemographic factors independently associated with unwillingness to vaccinate included younger age (<40 years) and attainment of less than a high school diploma. The effect of healthcare setting varied by ethnicity, with HCWs of Filipino ethnicity working in continuing care and HCWs of Caribbean ethnicity working in acute care having a greater likelihood of non-intent. Vaccination non-intent was associated with beliefs that i) vaccination was not required because of one’s own good health, ii) lower confidence that the COVID-19 vaccine would protect their family and patients, and iii) that getting vaccinated was not a professional responsibility. Vaccination non-intent was strongly associated with mistrust about how fast the vaccines were developed and vaccine safety concerns. Public health websites and healthcare providers were trusted sources of COVID-19 information. HCWs with vaccination intent were more likely to get vaccinated if direct financial supports (e.g. paid sick days) were provided (74% vs 25%, P<0.001).

**Table 2.**
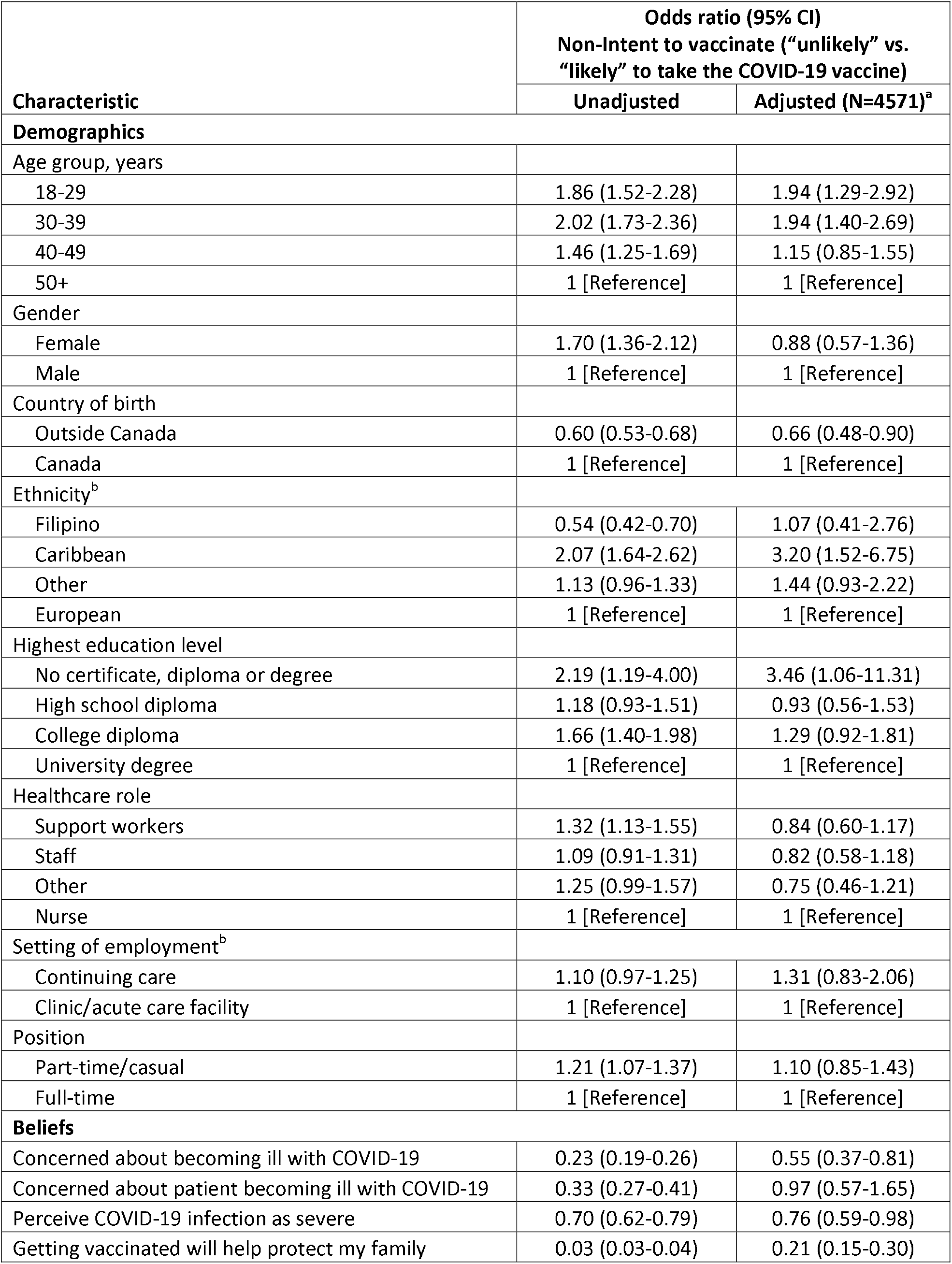
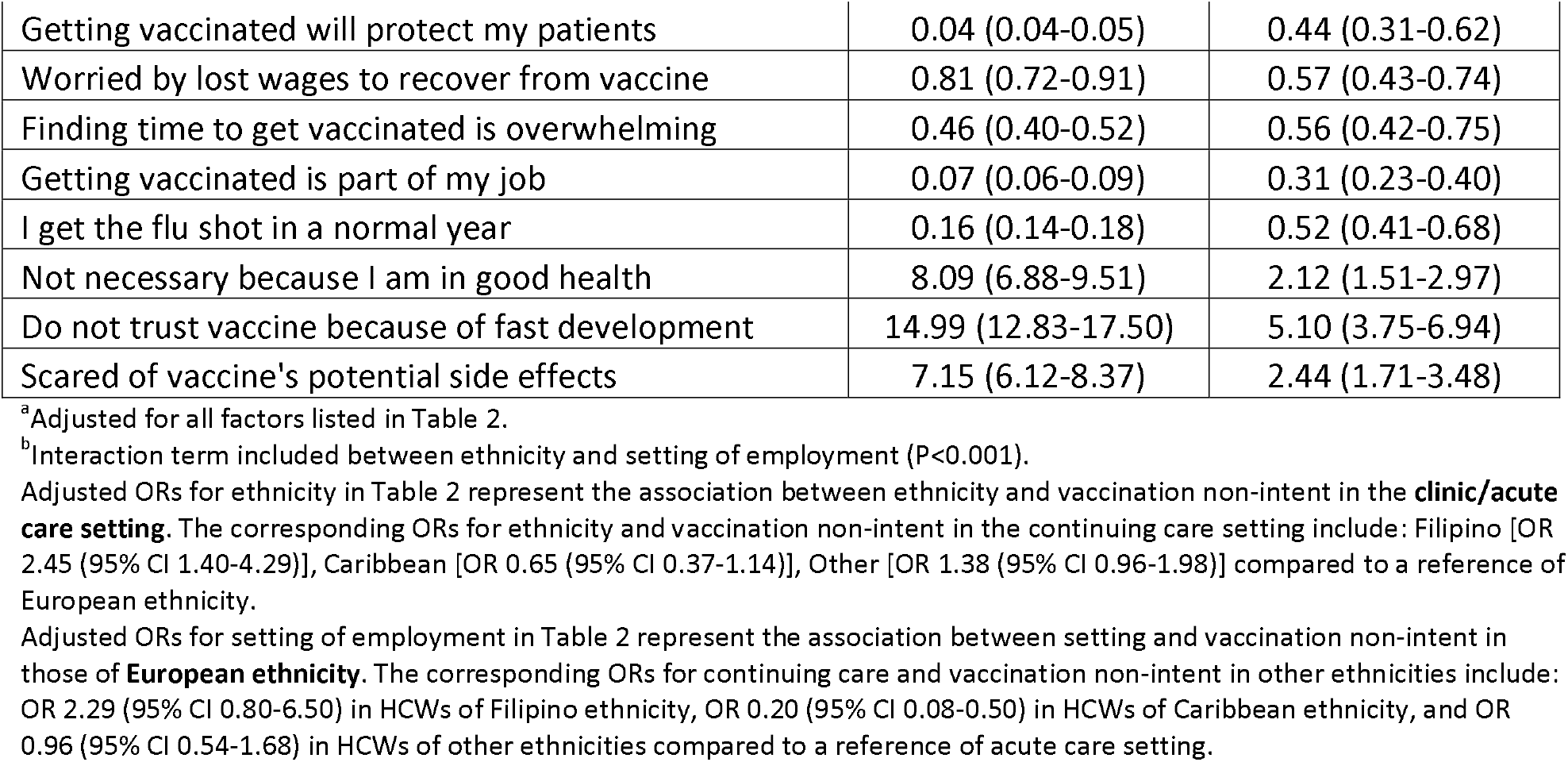
Factors associated with non-intent to take a COVID-19 vaccine.

## Discussion

This survey of 8,634 non-physician HCWs in Ontario, Canada indicated that the majority (80.4%) intend to take a COVID-19 vaccine. The 19.6% of non-physician HCWs expressing initial unwillingness to vaccinate are a key target for communication efforts and behaviorally informed strategies^4^, and may be underestimated due to social desirability bias. History of flu shot refusal may be a useful proxy for needing special outreach regarding COVID-19 vaccinations. Communication directed at non-physician HCWs^5^ should be tailored by ethnic subgroups and settings to increase salience. Messaging should leverage emotions (e.g., pride, hope, fear) to capture interest, while addressing safety concerns and confirming the low risk of side effects in contrast to the substantial morbidity and mortality of COVID-19. Emergent data about reduced transmission post-vaccination will be helpful. Communication channels should leverage credible sources including public health organizations, employers (i.e., healthcare institutions), and trusted healthcare providers.

In addition to targeted communication, financial and logistical supports (e.g., paid time off to get vaccinated, guaranteed sick leave in case of vaccine side effects, providing transportation to vaccine clinics or administering the vaccine on-site at workplaces) could further increase vaccine uptake, especially for this population who are often poorly paid without full-time work and benefits. ^6^

## Data Availability

The datasets generated during and/or analyzed during the current study are not publicly available as they are owned by the union who administered the survey.

## Acknowledgements

The authors would like to acknowledge the Service Employees International Union (SEIU), a union which represents over 2-million members across North America, and specifically SEIU Healthcare in Ontario for their partnership in designing and administering the survey. We would also like to acknowledge the healthcare workers who took the time to complete this survey and Dr. Mina Tadrous for his in-kind guidance regarding the analysis.

## Notes

### Competing Interest Statement

The authors have declared no competing interest.

### Funding Statement

No external funding was received for this work.

### Author Declarations

The study was approved by the Womens College Hospital Research Ethics Board.

### Summary of Updates

Corrected an error (belief associated with non-intent was that vaccination was *not* part of professional role) and added a co-author (who was previously in acknowledgements).

